# What Shapes HPV Vaccine Uptake Among Adolescent Girls from Urban Slums in Dhaka, Bangladesh: A Qualitative Study Using the WHO Behavioral and Social Drivers (BeSD) Framework

**DOI:** 10.64898/2026.08.14.26360437

**Authors:** Tonmoy Sarkar, Tamanna Sultana, Shayla Jesmin Nimmy, Md. Shariful Islam, Mohammad Ariful Islam, Farhat Jahan, Sazzad Hossain Khan, Kamal Ibne Amin Chowdhury, Md Tanvir Hossen, Md Abu Nayem, Nusrat Homaira, Farhana Haque, Abu Mohd Naser, A.S.M. Shahabuddin, S M Murshid Hasan, Saklayen Russel, Holly Seale, Firdausi Qadri, Syed Moinuddin Satter, Md Saiful Islam

**Author notes:** Corresponding author: Tonmoy Sarkar Research Investigator, icddr,b, Mohakhali, Dhaka 1212, Bangladesh.

## Abstract

**Background:** Human papillomavirus (HPV) is the leading cause of cervical cancer and the vaccine is the key preventive measure. In 2023, Bangladesh launched a school-based HPV vaccination campaign for girls aged 10-14 years. However, vaccine uptake among this group in urban settings remains suboptimal. This study explored adolescent girls’ (aged 10-14 years) understanding attitude, and motivation towards the vaccine, as well as the practical challenges impacting on vaccine uptake.

**Methods:** From April to June 2024, a qualitative study was undertaken in two urban slums in Dhaka, Bangladesh. Through a combination of convenience and snowball sampling, we conducted 15 in-depth interviews and one focus group discussion using the World Health Organization’s Behavioral and Social Drivers (BeSD) tool. Interviews were conducted in the native *Bengali* language, audio recorded, and transcribed verbatim. Framework analysis was performed to emerge key themes and generate study findings.

**Results:** A total of 26 girls with a mean age of 12.65 (SD: 1.23) participated in the study. While some participants believed that the HPV vaccine could reduce the infection during menstruation or prevent childbirth-related complications, there was uncertainty regarding the appropriate age for vaccination. Concerns were raised about menstrual irregularities, infertility, and the potential negative impact on marital prospects. Students spoke about being subjected to inappropriate jokes from their male peers. Male guardians were identified as the key decision makers and were perceived to be against the need for this vaccine. Operational barriers including inaccessible digital registration, limited information about the vaccine, and lack of systematic follow-up constrained the participation in the school-based HPV campaign.

**Conclusions:** Adolescents in urban slums faced multi-layered barriers, including knowledge gaps, cultural barriers, and accessibility challenges to HPV vaccination. Strengthening adolescent-friendly communication, engaging parents, teachers and male students, simplifying registration, adequate vaccine supply and ensuring supportive school-based vaccination processes are critical to improving equitable coverage and acceptance.

## Introduction

Human papillomavirus (HPV) infection is a major global public health challenge to women, particularly for cervical cancer [1, 2]. It is the second most common cancer in women, with 0.6 million new cases and around 0.35 million deaths each year [2]. Estimates from Global Cancer Statistics in 2018 indicate that most infections occur among young adults in their late teens and early twenties[1]. Almost 90% of total cases and deaths occur in low and middle-income countries (LMCIs) [1, 3]. Limited awareness, screening and lack of vaccination facilities are leading to at least six times higher age-standardized cervical cancer deaths in LMICs compared to high-income countries[4]. In Bangladesh, cervical cancer is the second most commonly diagnosed cancer, with approximately 12,000 new cases and more than 6,000 deaths reported each year [5]. More than 50 million women of reproductive age remain at risk of developing cancer, and projections suggest that without effective intervention, more than half a million Bangladeshi women will die from this disease by 2070 [6, 7]. Due to the lack of national cancer registries and restricted access to facilities, the true burden is underestimated [8]. Delayed diagnosis is another challenge that bounds the opportunities for treatment and contributes to the high fatality [8].

Vaccination against HPV infection has appeared as an essential approach for reducing cervical cancer. In high-income countries, where HPV vaccination coverage has reached around 80%, it has already proven to be an effective strategy. The World Health Organization (WHO) adopted a global strategy in 2020 to accelerate the elimination of cervical cancer, with a primary aim to vaccinating 90% of girls before the age of 15 by 20230 [9, 10]. In the line with the global initiative, the Government of Bangladesh launched the national HPV vaccination program in October 2023 for adolescent girls aged 10-14 years with the collaboration of WHO Bangladesh, Gavi, the Vaccine Alliance, and UNICEF Bangladesh [11, 12]. At the initial stage, the program was implemented for 18 days in the capital city, Dhaka, and gradually expanded to the other divisions across the country. Through this program, HPV vaccination was provided free of cost through educational institutions and designated vaccination centers. Students were able to receive this vaccine through an online registration process implemented as part of the program [11, 12]. While the reported national coverage for one dose vaccine reached 93% among eligible schoolgirls (aged 10-14) by December 2024, disparities remain in urban settings[13]. Although 66% of adolescent girls in urban Dhaka are attending school, only 38% of them received the vaccine in the school-based campaign. [13–15].

In Bangladesh, around 2.0 million adolescents live in informal settlements, and around 0.1 million of them reside in the slum areas of Dhaka city[13]. These urban slum areas are characterized by high population density, insecure housing, and limited access to basic health services. Adolescent girls from these areas are a particularly vulnerable subgroup as they have limited access to health literacy, accurate health information, reliance on informal social networks and continuous exposure to rumors [16]. Structural challenges such as poverty, complex health services, and limited access to digital devices further constrain participation in health programs [16, 17].

Moreover, limited studies explored the adolescent girl’s understanding of HPV infection and vaccination. Social and behavioral factors that influence vaccine uptake are mostly unrevealed. Addressing this limitation is crucial for finalizing the HPV vaccination strategy. This qualitative study aimed to explore the knowledge, perceptions, motivations, and practical challenges associated with HPV vaccination among adolescent girls living in urban slums in Dhaka, Bangladesh.

## Methods

### Study design

A cross-sectional study was conducted with a qualitative data collection approach. The study was guided by the Behavioral and Social Drivers (BeSD) framework of the World Health Organization[18].

### Study area

The study was implemented in two purposively selected slums in Dhaka, where icddr,b established health and demographic surveillance systems (HDSS) in 1998[19, 20]. These sites include the Korail and Kalshi slums in Dhaka. Korail slum spans approximately 0.4 square km, has around 200,000 inhabitants[19]. On the other hand, Kalshi slum consists of 0.25 square kmand about 30,000 people [20]. We considered several aspects for the selection process, including implementation of the initial vaccine campaign in the adjacent schools by the government in 2023, ensuring reliable information from the Health and Demographic Surveillance System (HDSS) platform and facilitating smooth entry in the community for the data collection (Figure 1).

**Figure 1.**
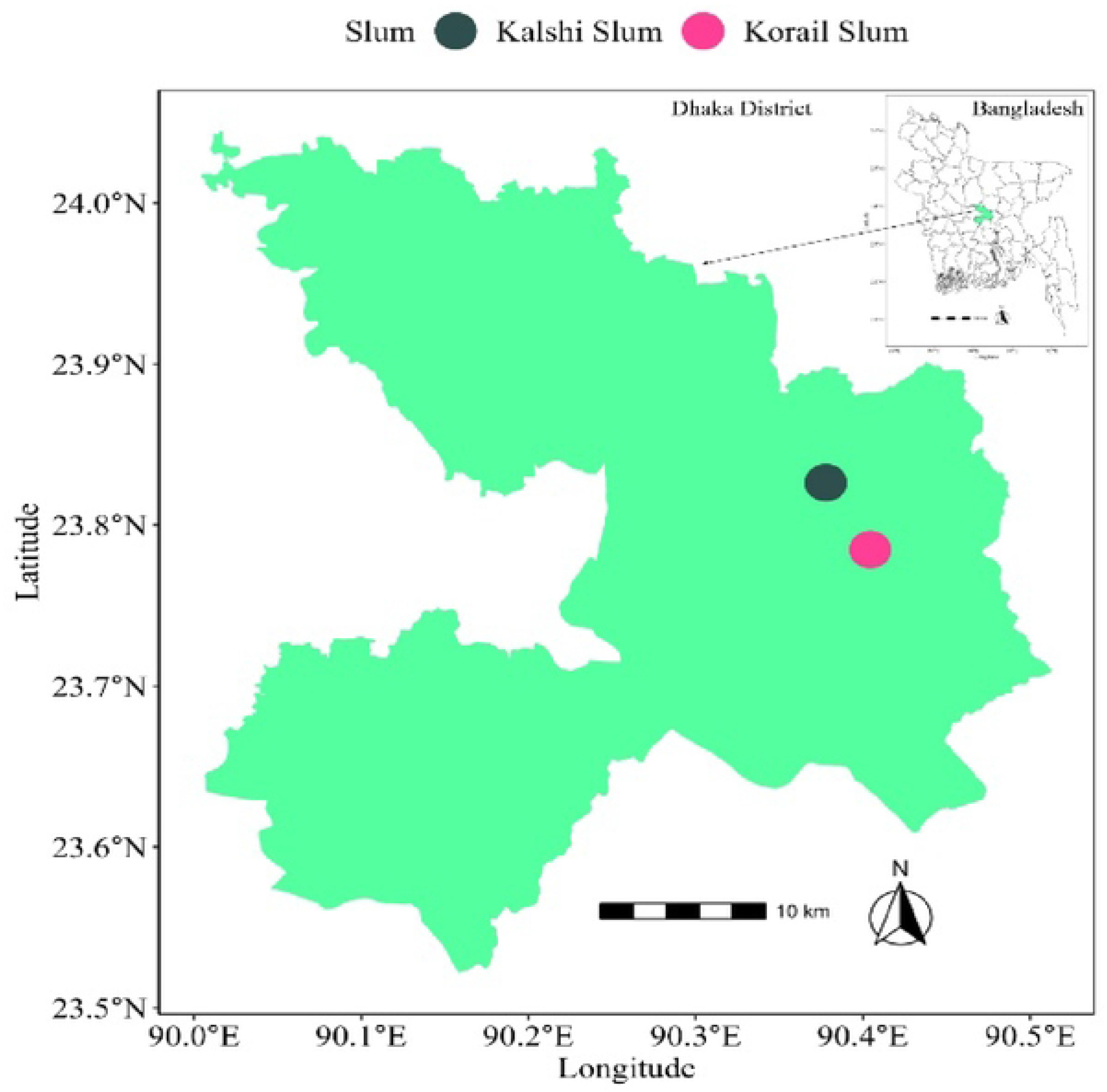
Study site

### Study population and sampling technique

Adolescent girls residing in selected slum areas were identified as the study population. The inclusion study criteria were as follows: (i) resident in the selected slums, (ii) enrolled in a school where the HPV vaccination campaign had been implemented, (iii) aged between 10 and 14 years, and (iv) provided informed assent and parental or guardian consent were eligible to participate in the study.

A combination of convenience and snowball sampling techniques was employed to recruit participants. An initial list of households with adolescent girls and their contact information was obtained from the HDSS, which has been operational since 1998. HDSS field staff served as the primary point of contact to identify potentially eligible households. The research team subsequently visited these households to verify the presence of adolescent girls within the eligible age range and to confirm the school enrolment status. The team also verified whether the HPV vaccination campaign had been implemented in the respective schools and assessed the vaccination status of potential participants.

For focus group discussions (FGDs), snowball sampling was used. Participants who had completed in-depth interviews (IDIs) were asked to refer peers within their social networks who met the eligibility criteria. The research team then contacted the referred individuals, provided detailed information about the study objectives and procedures, and invited them to participate in the FGDs.

## Data collection

Data collection was conducted between April and June 2024. In accordance with the qualitative guidance of the WHO Behavioral and Social Drivers (BeSD) framework, we developed semi-structured guidelines for IDIs and FGDs. The interview guidelines were organized around the four domains recommended by the WHO tool: thinking and feeling, social processes, motivation, and practical issues (Supplementary Table 1).

To ensure relevance, clarity, and contextual appropriateness, the instruments were pilot tested with adolescent girls (aged 10-14 years) residing in a separate slum area at Mohakhali in Dhaka city. Based on feedback and field observations from the pilot phase, the guidelines were refined and finalized.

A team of three trained female social scientists with extensive experience in qualitative research conducted data collection activities. Both vaccinated and non-vaccinated adolescent girls were enrolled as study participants. Each in-depth interview lasted approximately 45 to 75 minutes, while the FGD lasted around 80 minutes. Interviews were conducted in a private, secure settings to foster a supportive environment and enable participants to share their views openly and without fear.

## Data analysis

We conducted a framework analysis to organize the findings under the four domains of the BeSD tool (Supplementary Table 1). In-depth interviews (IDIs) and focus group discussions (FGDs) were audio-recorded using digital recorders, anonymized with non-identifying codes, and securely stored in cloud-based systems. Informal conversations were expanded into detailed field notes. All recordings were transcribed verbatim and translated from Bangla into English.

Two team members independently reviewed a subset of transcripts and field notes to develop an initial coding framework. The research team then coded all transcripts manually, refining and expanding codes iteratively as new themes emerged. Discrepancies or new codes were discussed among the team members to reach consensus, ensuring consistency and intercoder reliability.

Codes were subsequently categorized and mapped to the four BeSD domains to analyse knowledge, perceptions, attitudes, and experiences related to HPV vaccination.

## Ethics statement

The study protocol was approved by the institutional review board of icddr,b. Informed written assent was obtained from the study participants and also written consents were obtained from the legal guardians. All the recordings were handled confidentially and personal identifiers were removed or anonymized in the analysis.

## Result

### Demographic characteristics of the study participants

A total of 26 adolescent girls participated in the study, with an average age of 12.65 (±1.23) years. All the participants were students and unmarried. More than half of the participants (54%, n=14) were studying at junior level (Class six-eight), and the rest were enrolled from higher secondary level (Class nine-ten). On the other hand, 54% (n=14) of participants received the HPV vaccine during the school campaign program. Nearly half of the participants (46%, n=12) reported family income of BDT 20,000-30,000 (approximately USD 167-250). Around 23% (n=6) had a monthly income of BDT 10000-20000 (USD 83-167), while 15% each belonged to the household earning ≤BDT 10,000 (≤USD 83) and ≥30,000 (≥USD 250). In terms of family size, most participants belonged to households with 5-10 family members (62%, n=16) followed by households with fewer than 5 members (35%, n=9). Only one participant reported living in a household with more than 10 members.

### Thinking and feeling

Questions were asked about participants’ knowledge and perceptions regarding cervical cancer. The majority (n=9) of the vaccinated adolescent girls described cervical cancer as a life-threatening contagious disease for women of reproductive age that was linked to the HPV infection. A few participants (n=4) believed that poor menstrual hygiene practices, such as using contaminated or old pads and having multiple pregnancies, were major causes of this infection. One participant stated that surgical treatment could prevent further progression of the cervical cancer. However, some participants from both IDIs (n=5) and FGD (n=1) emphasized that immunization was essential for preventing the future occurrence of the disease. Of those who reported having some knowledge such as information about HPV and the vulnerability of female students, they received it from school teachers, family elders and from social media. One vaccinated student from class seven expressed that,

> “This cancer is easily spreadable to the whole body from cervix. Though it’s a cancer, it’s really dangerous and worldwide, many females die. Adolescent girls are more vulnerable for cervical cancer. Sometimes surgery can prevent the death however it does not cure.”

One of the participants mentioned that HPV vaccine could boost the immune system among adolescent girls, helping the body fight against the virus. The majority of the participants also believed that this vaccine prevented uterine tumors and also protected against cancer in other parts of the body. Additionally, they perceived that it reduced the risk of infection during menstruation and childbirth-related complications. One vaccinated participant said that,

> “We, the girls, have to face many health challenges during menstruation, including fever, headache, reduced physical movement, and abdominal pain. The vaccine can help protect against the germs that enter the body during menstruation, ultimately preventing the future occurrence of cervical cancer.”

Most of the participants told that adolescents aged 10-14 years were eligible for the HPV vaccine. Some stated (n=3) that women of reproductive age could receive this vaccine at any time during their reproductive years. One participant noted that women with multiple pregnancies might have a greater need for this vaccine. She reported that one elderly neighbor expressed concerns and shared her opinion regarding the importance of vaccine as:

> “We had multiple pregnancies with short spacing. Something is coming out of the vagina and feels wet most of the time. This provides an opportunity to get infected, which ultimately turns into cancer. This vaccine can prevent this infection the vaginal infection.”

Half of the vaccinated study participants (50%) commonly reported mild side effects after vaccination, including fever, headache, and mild to moderate pain at the injection site on the upper arm. One individual raised concerns that the HPV vaccine might affect girls’ menstrual cycle, which she believed could negatively impact future reproductive health and potentially lead to marital conflict or even divorce. One study participant told that,

> “Girls experience natural health issues such as the menstrual cycle. This vaccine may disrupt that regularity. Later in life, we need to marry and have children, and if the menstrual cycle is affected, we may face difficulties in conceiving, which would be painful. As a result, this could even lead to divorce.”

### Motivation

During the school-based HPV vaccination campaign, adolescent girls mainly depended on their legal guardians’ and school teachers’ opinions for vaccination. After the initial launch of the vaccine program through an educational institute-based campaign, many guardians felt anxiety and doubt about the vaccine. Few participants (approximately 12%) reported that lack of knowledge, limited awareness, and challenges to accessing proper information likely contributed to this hesitancy. Moreover, doubtful conversations about the effectiveness of the HPV vaccine with peer groups in the slum settlement also spread uncertainty among the male guardians.

Before vaccination, adolescent participants primarily feared the administration process of the vaccine, particularly the use of a needle. One of the study participants who did not receive the vaccine said that,

> “One of my neighboring uncles asked my father why you are intending to provide this vaccine to your daughter? This vaccine will not make any changes to your daughter’s health. These are not effective. I think you should wait few days and then make a decision about providing this vaccine.”

Some guardians were hesitant due to a lack of trust in the health system and concern about the cost and effectiveness of the vaccine. They believed that a vaccine provided free of charge by the government might be less effective for a severe and costly disease like cervical cancer.

Moreover, the number of required HPV vaccine doses also triggered confusion among participants as they noticed a difference between the two-dose schedule used in private settings and the single dose provided by the government at the school campaign. One participant who received an HPV vaccine said that,

> “We received one dose HPV vaccine during our school campaign. As far as I know, those who purchased it privately must take two doses. From the school authority, we did not hear anything for the next doses. We are waiting for that, and see what they say about it. We don’t know this one dose will be how much effective.”

Some participants emphasized in receiving the vaccine through the school campaign, as this vaccine was not available in all the vaccine centers or health facilities. One study participant said that,

> “Initially, I was afraid and wondered what might happen to me if I did not take the vaccine. I discussed it with my teachers and friends. Many of my friends told me that since the government has initiated this vaccine, it must be for our benefit. Moreover, my mother said, school is arranging the campaign so you should take this. I do not want you to suffer from this disease in the future.”

### Social processes

According to the study participants, most were motivated to receive the HPV vaccine through female guardians such as their mother, elder sister, or aunt. Classroom discussions led by female teachers played a vital role in shaping their understanding and motivation. Many adolescents felt uncomfortable discussing cervical cancer and vaccination in group settings, particularly in front of their fathers or male family members, as well as in front of male peers at school. Some participants cited healthcare workers, peer influence, and self-learning through social media as sources of motivation. One of the study participants said that,

> “After hearing the announcement of the vaccination campaign from the school, I informed my mother. I did not have any discussion with my father as I felt shy to discuss this matter with him. He stays at home and I think he has a good understanding of the matter. After hearing from me, my mother talked with my father, and he was initially discouraged in receiving the vaccine.”

Multiple rumors were reported by the participants, including that water would be used instead of real vaccines, and viruses could be transmitted through reused needles. A non-vaccinated student from class eight stated:

> “No need to take the vaccine from here as they vaccinate with the same needle… they said, as they vaccinate many people with the same needle, many viruses can spread as a result… yes, they said, they will vaccinate others there with one needle. As the government is providing free of cost, they are vaccinating all with one needle. They might doubt the standard of the vaccine.”

Participants also reported being blamed and stigmatized by the neighborhood and male peers. Some of them experienced dirty jokes by their male peers after receiving the vaccine, which shaped the vaccine decision and also discouraged school attendance following vaccination.

### Practical challenges

Participants reported that a digital birth certificate was required to complete the online registration process to receive the vaccine. The lack of digitalization of the birth certificate, limited access to smartphones or laptops, lack of skill for self-registration, and difficulties in accessing the online portal both in cyber cafes as well as school-based registration, created substantial barriers among slum dwellers. One study participant said that,

> “Not all guardians had the same level of awareness, and many did not have the skill for the self-registration process. They need support. Many families did not have access to Android phones. They had to complete the registration process through local cyber cafes. There, they faced challenges in accessing the registration portal. Some students asked how I completed my registration process. I told them that I finished my registration at night, especially after 12:00 am.”

The short notice during the implementation of the school-based vaccination campaign contributed to further registration-related challenges. Several students reported difficulties with age verification due to inaccuracies in digital birth certificates, including incorrect dates of birth. In addition, participants described vaccine shortages and a limited number of vaccination sites during the campaign period.

Concerns were also raised regarding the availability of adequately trained vaccinators and the limited psychological support provided by school authorities. Observing bleeding at the injection site on the upper arm among some vaccine recipients increased fear and anxiety. Participants further noted that schools did not provide sufficient information about alternative vaccination centres when doses were unavailable on site, nor did they clearly communicate available support for managing post vaccination adverse events.

Moreover, some participants reported the absence of prior discussions involving teachers and guardians within the school setting. Insufficient oversight and follow-up by school authorities reportedly resulted in some eligible students being missed during the campaign, contributing to uncertainty and confusion among adolescents.

## Discussion

Our study highlights important gaps in adolescent girls’ understanding, their perception and motivation about the HPV vaccine among those living in two urban informal settlements of Dhaka. Although a majority of participants demonstrated some awareness of the HPV vaccine and its role in preventing cervical cancer, their motivation was challenged and shaped by a wide range of misinformation. Many participants expressed a lack of knowledge on infection and its transmission also shaped their motivation regarding the HPV vaccine decision. Despite misunderstanding, participants shared positive thoughts about HPV vaccination and recognized its preventive benefits. Concerns about potential long-term effects related to the menstrual cycle and future infertility further influenced vaccine hesitancy and uptake. Multiple operational challenges within the school-based campaign, including short-notice scheduling, inadequate registration support, and limited vaccination counseling, also contributed to vaccine decision.

Our study findings emphasize the importance of continuing to educate adolescent girls and their relevant stakeholders. Parents, school teachers, and health professionals should be included in this continuous awareness activity to promote vaccine confidence. Several studies raised concerns about the safety and efficacy of the HPV vaccine, which continued to be a barrier to the vaccination campaigns [21–23]. On the other hand, trust in vaccine efficacy and safety may play a role in vaccination [21, 23]. Although study participants reported various temporary side effects, some revealed concern related to the long-term consequences of their reproductive capacity. The association between HPV vaccination and perceived menstrual irregularities has already been highlighted in prior literature [24, 25]. These results emphasized that gender norms and reproductive expectations shape vaccine decisions among adolescents. Robust clinical and behavioral research is required for better illustration of whether such adverse effects are biologically plausible or primarily driven by misinformation, rumors, and community beliefs.

The HPV vaccine is closely associated with reproductive and sexual health, which remains culturally sensitive in the context of Bangladesh[26]. Our study revealed that this sensitivity restricts open communication among adolescents and their male guardians or teachers. As a result, they often rely on peers which often reinforces misconceptions and incomplete knowledge. In addition, household decision-making dynamics affected vaccination outcomes. Adolescents girls from the informal settlement usually depend on their fathers’ decisions for accessing any health services[17, 27]. Though mothers played a central role in guiding decisions, participants reported disagreements within families. Some parents encouraged vaccination based on health benefits, while others discouraged it due to fears of social stigma and circulating rumors. The inconsistent guidance limited adolescents’ autonomy and delayed the timely uptake of the vaccine. Strengthening parents’ involvement through slum-based awareness sessions may help counter misconceptions and improve vaccine confidence.

Rumors emerged as a powerful influence in shaping vaccine decisions[28]. Widespread rumors, such as the reuse of needles and the administration of water instead of vaccine reagents, created substantial concern among adolescents as well as their guardians. These narratives spread rapidly through informal social networks where health literacy is low. Even after getting the accurate information from the school teachers and mothers, many lacked structured and appropriate messages about the HPV vaccine. Social stigma added another layer of complexity for vaccine decision. Some adolescent girls experienced teasing, shame, or inappropriate remarks from male peers at their schools. These attitudes encountered open discussions in the classroom and influenced the vaccine uptake[29, 30]. Study findings underscore the need for adolescent-friendly communication that may foster a supportive environment for open discussion and reduce social stigma [29].

Studies from different settings have shown that higher vaccine prices are associated with lower uptake. In our context, the provision of a free vaccine raised concern among recipients regarding its quality and efficacy [31, 32]. Some guardians perceived government-provided free vaccines to be of lower quality or potentially unsafe, which ultimately reduced the acceptance. Our study also found that the number of required HPV vaccine doses played a critical role in shaping the perception of vaccine efficacy. On the other hand, students were unsure why the school campaign offered a single dose while private providers recommended two doses, leading to concern about the vaccine completion and its efficacy. However, trust improved notably when educational institutions endorsed the campaign activities. This emphasized the value of institutional collaboration in strengthening vaccine credibility[33, 34].

Operational challenges of the school-based HPV campaign also influenced vaccine perception among slum adolescents[35]. Short-notice scheduling of the campaign provided limited time for preparation of school authorities, guardians, and students for full participation. Unavailability of the digital birth certificate, missed registration deadline, and student absences on vaccination day reduced overall coverage. These barriers may have affected adolescent girls from informal settlements, particularly those who were not regularly attending school or had already dropped out. School-based campaigning is an effective strategy for reaching a large number of adolescents. In the context of informal settlers, complementary community-based approaches are needed to ensure equitable access. Outreach through community health workers, slum-based awareness activities, targeted social media campaigns, and catch-up vaccination programs could help identify and vaccinate out-of-school girls who may otherwise be missed. Simplifying the registration process, such as allowing offline/assisted enrollment, and ensuring flexible verification systems can reduce missed vaccinations[31]. Advanced coordination between the health and education sectors and families can simplify this registration process and track all eligible adolescents.

Our study revealed that fear of injections, hurried administration, and insufficient information before and after vaccination created discomfort and negative impressions among adolescents. Several eligible students missed the vaccine due to the weak monitoring system from the school authority. The absence of mechanisms for student follow-up, management of adverse events following immunization, or alternative vaccination points contributed to uncertainty and reduced the overall effectiveness of the program [36]. Schools require timely teacher training, clear communication about dose schedules, and preparedness to manage students’ fear of injections.

Psychological support before vaccination and improved monitoring can reduce confusion and missed opportunities.

## Limitation

Participants were recruited from selected urban slum communities in Dhaka. Findings from this study may not be transferable to rural populations, non-slum urban residents, or other regions of Bangladesh. Urban slum settings are characterized by unique socioeconomic vulnerabilities, mobility patterns, and health service access dynamics that may shape vaccine decisions, which are diverse from other contexts. The use of purposive sampling may have resulted in the under- representation of families who are less engaged with schools, socially isolated, or highly vaccine-hesitant. Social desirability bias may have influenced responses. Given that vaccination is widely promoted through government and public health messaging, participants may have felt pressure to express non-supportive attitudes toward the vaccine. This concern is particularly relevant in focus group discussions, where peer influence and dominant voices may have shaped group narratives. Additionally, given the age range of adolescents (10–14 years), participants may have deferred to parental views or felt uncomfortable with discussion topics that indirectly linked to sexual and reproductive health. Although the BeSD framework provided a structured lens for data collection and analysis, reliance on this framework may have limited exploration of broader structural and political determinants of vaccine uptake. Factors such as extreme poverty, informal settlement precarity, and systemic mistrust may extend beyond the core BeSD domains and warrant further investigation.

## Conclusion

Our study demonstrates that adolescent girls living in urban informal settlements encountered multiple challenges to HPV vaccination. Knowledge gaps, pervasive misinformation, socio- cultural norms, and operational challenges were widely reported by study participants. Although the preventive benefits of HPV vaccine were reported, however concern about side effect, rumors, social stigma, and inconsistent household guidance contributed to vaccine hesitancy.

Operational constraints such as short-notice campaign arrangement, digital registration difficulties, inadequate vaccine supply, and limited support for the post-vaccination period further influenced vaccine decisions. These findings underscore the importance of tailored adolescent-friendly health education and the active engagement of parents, teachers and health professionals. Strengthened coordination between education and health authorities is also essential to improve vaccine confidence, reduce misconceptions, and ensure equitable vaccine coverage in the marginalized urban population.

## Data Availability

The data underlying the findings of this study consist of qualitative interview and focus group transcripts containing potentially identifiable information from adolescent participants. To protect participant confidentiality and comply with the ethical approval and informed consent procedures, these data are not publicly available. De-identified excerpts supporting the findings are included within the manuscript. Additional de-identified data may be made available upon reasonable request to the corresponding author, subject to approval by the Institutional Review Board (IRB) of icddr,b and in accordance with applicable ethical and legal requirements to ensure participant privacy.

## Acknowledgements

We are grateful to the government of Bangladesh and Canada for providing core/unrestricted support to the icddr,b the home institution of the primary author. This study was funded by the Sabin Vaccine Institute, whose support made this research possible. We extend our sincere gratitude to all study participants, as well as local administration for their time and cooperation.

## Declaration of generative AI and AI-assisted technologies in the writing process

During the preparation of this work, we used ChatGPT AI to assist with grammar checking and language refinement. After using this tool, the corresponding author and senior author reviewed and edited the content as needed and take full responsibility for the final content of the publication.

## References

1. Sung, H., et al., Global Cancer Statistics 2020: GLOBOCAN Estimates of Incidence and Mortality Worldwide for 36 Cancers in 185 Countries. CA Cancer J Clin, 2021. 71(3): p. 209–249.

2. Organization, W.H. Cervical cancer. 2024; Available from: https://www.who.int/news-room/fact-sheets/detail/cervical-cancer.

3. McDowell, S. Cervical Cancer Leads Cancer Deaths for Women in 37 Countries. 2024; Available from: https://www.cancer.org/research/acs-research-news/cervical-cancer-leads-cancer-deaths-37-countries.html.

4. Singh, D., et al., Global estimates of incidence and mortality of cervical cancer in 2020: a baseline analysis of the WHO Global Cervical Cancer Elimination Initiative. The Lancet Global Health, 2023. 11(2): p. e197–e206.

5. Banik, R., et al., Investigating Bangladeshi Rural Women’s Awareness and Knowledge of Cervical Cancer and Attitude Towards HPV Vaccination: a Community-Based Cross-Sectional Analysis. J Cancer Educ, 2022. 37(2): p. 449–460.

6. Bruni L, A.G., Serrano B, Mena M, Collado JJ, Gómez D, Muñoz J, Bosch FX, de Sanjosé S. and ICO, Human Papillomavirus and Related Diseases Report. 2023.

7. Uddin, A., et al., Cervical Cancer in Bangladesh. South Asian J Cancer, 2023. 12(1): p. 36–38.

8. Denny, L., M. Quinn, and R. Sankaranarayanan, Chapter 8: Screening for cervical cancer in developing countries. Vaccine, 2006. 24 Suppl 3: p. S3/71–7.

9. Ghebreyesus, D.T.A., Global strategy to accelerate the elimination of cervical cancer as a public health problem 2020, World Health Organization

10. Garland, S.M., et al., IPVS statement on “Temporary HPV vaccine shortage: Implications globally to achieve equity”. Papillomavirus Research, 2020. 9: p. 100195.

11. Organization, W.H. Human Papillomavirus (HPV) Vaccination Launching in Bangladesh: A single dose vaccine has potential to prevent cervical cancer! 2023; Available from: https://www.who.int/bangladesh/news/detail/10-10-2023-human-papillomavirus-(hpv)-vaccination-launching-in-bangladesh--a-single-dose-vaccine-has-potential-to-prevent-cervical-cancer.

12. Bangladesh, U. The Interim Government of Bangladesh launches the final phase of the human papillomavirus (HPV) vaccination campaign, targeting over 6.2 million girls, to achieve nationwide coverage. 2024; Available from: https://www.unicef.org/bangladesh/en/press-releases/interim-government-bangladesh-launches-final-phase-human-papillomavirus-hpv.

13. Maria Quattria and K.W., Child labour and education – A survey of slum settlements in Dhaka (Bangladesh). World Developement Perspective 2019.

14. Mohammad, A.A. Bangladesh makes HPV vaccine routine after successful launch campaign. 2025; Available from: https://www.gavi.org/vaccineswork/national-hpv-vaccine-programme-goes-routine-bangladesh-can-boast-93-coverage-rate.

15. Alaml, A., et al., HPV VACCINE COVERAGE AMONG ADOLESCENT GIRLS FOLLOWING A NATIONAL VACCINATION PROGRAM: A COMMUNITY-BASED CROSS-SECTIONAL STUDY IN BANGLADESH. 2025.

16. Hasan, M.Z., et al., Assessment of socioeconomic and health vulnerability among urban slum dwellers in Bangladesh: a cross-sectional study. BMC Public Health, 2024. 24(1): p. 2946.

17. Van der Heijden, J., et al., ’Working to stay healthy’, health-seeking behaviour in Bangladesh’s urban slums: a qualitative study. BMC Public Health, 2019. 19(1): p. 600.

18. Organization, W.H. Behavioural and social drivers of vaccination: tools and practical guidance for achieving high uptake. 2022; Available from: https://www.who.int/publications/i/item/9789240049680.

19. Rahman, M.M., et al., Differences in access to water, sanitation, and hygiene facilities among residents of Korail Slum, Bangladesh, during normal vs. water-logging situations. PLoS One, 2025. 20(9): p. e0332534.

20. Razzaque, A., et al., Cohort Profile: Urban Health and Demographic Surveillance System in slums of Dhaka (North and South) and Gazipur City Corporations, Bangladesh. International Journal of Epidemiology, 2023. 52(5): p. e283–e291.

21. Zheng, L., J. Wu, and M. Zheng, Barriers to and Facilitators of Human Papillomavirus Vaccination Among People Aged 9 to 26 Years: A Systematic Review. Sex Transm Dis, 2021. 48(12): p. e255–e262.

22. O. Ricke, D., Menstrual Adverse Events Post COVID-19 and HPV Immunization. Preprints, 2025.

23. Santos, S.A.D., et al., Comparison between the safety of the HPV vaccine versus placebo: a systematic review and meta-analysis of randomized clinical trials. J Pediatr (Rio J), 2025. 101(5): p. 101411.

24. Wastila, L., et al., Association Between Vaccination for Human Papillomavirus (HPV) and Autonomic Dysfunction and Menstrual Irregularities: A Self-Controlled Case Series Analysis. Drugs Real World Outcomes, 2025. 12(3): p. 467–477.

25. Gong, L., et al., Human papillomavirus vaccine-associated premature ovarian insufficiency and related adverse events: data mining of Vaccine Adverse Event Reporting System. Sci Rep, 2020. 10(1): p. 10762.

26. Sultana, S., et al., Knowledge and willingness towards human Papillomavirus vaccination among the parents and school teachers of eligible girls in Dhaka, Bangladesh: A school-based cross- sectional study. J Virus Erad, 2025. 11(1): p. 100590.

27. Haque, M.A., et al., Social norms and maternal health information-seeking behavior among adolescent girls: A qualitative study in a slum of Bangladesh. PLoS One, 2024. 19(12): p. e0315002.

28. Islam, M.S., et al., COVID-19 vaccine rumors and conspiracy theories: The need for cognitive inoculation against misinformation to improve vaccine adherence. PLoS One, 2021. 16(5): p. e0251605.

29. McKenzie, A.H., et al., Parents’ stigmatizing beliefs about the HPV vaccine and their association with information seeking behavior and vaccination communication behaviors. Hum Vaccin Immunother, 2023. 19(1): p. 2214054.

30. Wong, L.P., et al., Multidimensional social and cultural norms influencing HPV vaccine hesitancy in Asia. Hum Vaccin Immunother, 2020. 16(7): p. 1611–1622.

31. Xu, M.A., et al., Improving HPV Vaccination Uptake Among Adolescents in Low Resource Settings: Sociocultural and Socioeconomic Barriers and Facilitators. Adolesc Health Med Ther, 2024. 15: p. 73–82.

32. Wang, W., The impact of vaccine access difficulties on HPV vaccine intention and uptake among female university students in China. Int J Equity Health, 2025. 24(1): p. 4.

33. Perlman, S., et al., Knowledge and awareness of HPV vaccine and acceptability to vaccinate in sub-Saharan Africa: a systematic review. PLoS One, 2014. 9(3): p. e90912.

34. Chan, D.N.S., et al., Factors affecting HPV vaccine uptake among ethnic minority adolescent girls: A systematic review and meta-analysis. Asia Pac J Oncol Nurs, 2023. 10(9): p. 100279.

35. Siu, J.Y., A. Lee, and P.K.S. Chan, Schoolteachers’ experiences of implementing school-based vaccination programs against human papillomavirus in a Chinese community: a qualitative study. BMC Public Health, 2019. 19(1): p. 1514.

36. Davies, C., et al., Effect of a School-Based Educational Intervention About the Human Papillomavirus Vaccine on Psychosocial Outcomes Among Adolescents: Analysis of Secondary Outcomes of a Cluster Randomized Trial. JAMA Netw Open, 2021. 4(11): p. e2129057.

